# Clinical phenotypes in hypertension: a data-driven approach to risk stratification and outcome prediction

**DOI:** 10.1101/2025.04.22.25326237

**Authors:** Elisa Rauseo, Ahmed M. Salih, Jackie Cooper, Musa Abdulkareem, Christopher R.S. Banerji, Sucharitha Chadalavada, Hafiz Naderi, Patricia B Munroe, Anthony Mathur, Nay Aung, Gregory G. Slabaugh, Steffen E. Petersen

## Abstract

**Background:** Hypertension (HTN) is a major contributor to cardiovascular (CV) morbidity and mortality. Its heterogeneity complicates risk stratification. Unsupervised machine learning can identify risk profiles and refine preventative strategies.

**Objectives:** This study applies clustering analysis to clinical data to identify HTN phenogroups and their link with CV abnormalities and outcomes.

**Methods:** 14,840 UK Biobank participants with a diagnosis of HTN who underwent cardiovascular magnetic resonance (CMR) were analysed. K-means clustering was applied to 77 clinical variables. Associations with incident HF, atrial fibrillation (AF), atherosclerotic events, all-cause mortality, and major adverse cardiovascular events (MACE) were examined. Mediation analysis assessed the role of CV imaging metrics in risk stratification.

**Results:** Three clusters emerged. Cluster 1, predominantly female with the most favourable metabolic profile, had the lowest risk. Cluster 2, predominantly male with the highest atherosclerosis burden, carried the greatest risk, particularly for AF and HF (Hazard ratio [HR] 1.80 and 1.85; p <0.005). This group showed the most severe cardiac remodelling, impaired cardiac mechanisms, and global left atrial (LA) dysfunction, approaching the HF myocardial substrate. Cluster 3 had an intermediate risk profile resembling metabolic syndrome, with moderate MACE risk and a higher susceptibility to AF (HR 1.30 and 1.61; p <0.05). While in cluster 2 the risk was largely mediated by LV remodelling, in cluster 3, its role was attenuated and more evenly balanced with LA dysfunction.

**Conclusions:** Clustering analysis revealed distinct HTN phenotypes with unique clinical and imaging profiles, enhancing risk stratification and supporting more individualised treatment strategies.

## Introduction

Essential hypertension (HTN) is a major modifiable risk factor contributing to global mortality and morbidity (1). Chronic HTN can impact target organs, leading to structural and functional changes in the heart, arteries, kidneys, and brain. Without treatment, these changes may worsen, resulting in severe complications such as heart failure (HF), myocardial infarction (MI), atrial fibrillation (AF) stroke, cognitive decline, and kidney diseases. These chronic conditions can significantly impact patients’ outcomes, highlighting the importance of effective HTN risk assessment and preventive measures (1).

While blood pressure reduction remains central to HTN management, multiple factors influence disease trajectory, treatment response, and outcomes. These include environment, socioeconomic, psychological, demographic, lifestyle, genetic, and systemic mechanisms across multiple organs. This heterogeneity complicates risk stratification and management. Traditional risk stratification models rely largely on blood pressure thresholds and traditional risk factors to predict 10-year cardiovascular disease risk (1). Although guidelines now recommend integrating non-traditional modifiers (e.g. socioeconomic status) to refine risk assessment and guide management, these approaches fail to capture HTN heterogeneity, leaving some patients at high risk despite optimal blood pressure control, while others experience adverse outcomes even at lower blood pressure levels (1,2). This underscores the need for a more comprehensive approach that accounts for the full spectrum of contributing factors and enables personalised risk-based management.

Clustering is an unsupervised machine-learning method that identifies patterns and subgroups within heterogeneous populations. It has improved phenotyping and risk stratification for several cardiovascular diseases, including HF, and provided pathophysiological insights (3,4).

Previous clustering studies in HTN have identified clinically significant phenotypes, broadening classification beyond blood pressure values and offering further insights for guiding personalised preventative measures treatments (5–9). However, most previous studies had limited sample sizes, few clinical variables for risk stratification, and lacked links between clinical phenotypes, cardiovascular (CV) imaging changes, and long-term outcomes.

This study utilises clustering analysis on a large UK Biobank cohort to refine risk stratification in hypertensive patients and address its inherent challenges. By identifying clinical phenotypes from multidimensional clinical data, and linking them to CV imaging and outcomes, it aims to uncover distinct risk mechanisms. This unbiased approach may offer a more refined risk assessment and support tailored treatment strategies to improve clinical outcomes.

## Methods

### The UK Biobank

The UK Biobank is a large prospective cohort study collecting detailed phenotypic and genotypic information from over 500,000 participants across the UK (10). Baseline information includes socio-demographics, lifestyle, medical history, genetics, physical measures, and health-related outcomes. Cardiovascular magnetic resonance (CMR) imaging was conducted on more than 50,000 participants.

This study complies with the Declaration of Helsinki and received ethical approval from the NHS National Research Ethics Service (17 June 2011, Ref 11/NW/0382; extended on 18th June 2021, Ref 21/NW/0157). All participants provided written informed consent.

### Study population

The study population included UK Biobank participants with a confirmed diagnosis of HTN, defined through a combination of ICD codes, self-reported disease, doctor-diagnosed conditions, and the use of antihypertensive medications. Blood pressure values alone were not used to define HTN status, as isolated elevations may not reflect established disease. To focus on individuals with HTN without advanced cardiac pathology, we excluded those with a history of HF at the time of imaging. This approach aimed to capture a clinically relevant HTN cohort, free from overt HF, to explore phenotypic differences and their association with cardiovascular risk. The specific UK Biobank field codes used to define the study population are detailed in Supplemental Table 1.

### Phenotypic domains

Baseline characteristics of the study population, including socio-demographic factors, lifestyle habits, physical attributes, ECG parameters, and laboratory data, were collected and used as phenotypic domains for clustering analysis. The full list of phenotypic features scrutinised for clustering, with corresponding UK Biobank data fields, is reported in Supplemental Table 2. A more detailed description of how the phenotypic features were calculated is provided in a previous publication (3).

### Imaging parameters

CMR-derived features, not included in clustering analysis, were utilised in *post hoc* analyses to validate clustering results and assess their impact on outcomes. A detailed description of the CMR protocol and image analysis is provided in the Supplemental Materials. Briefly, CMR images were acquired with a 1.5 Tesla scanner (MAGNETOM Aera, Siemens Healthcare) using a previously described protocol (11). Image analysis used Circle’s CVI42 software (Circle Cardiovascular Imaging Inc.), with metrics derived from an automated tool with inbuilt quality control.

This study included several CMR measures, which were indexed for body surface area, where applicable. The following metrics were used: left and right ventricular (LV, RV) volumes and stroke volumes; LV mass; LV maximum wall thickness; LV mass-to-volume (M/V) ratio; ejection fractions for both ventricles; LV global function index; LV myocardial native T1 measured in mid-short-axis; cardiac mechanics indices, such as global longitudinal, circumferential, and radial strain (GLS, GCS, GRS), and torsion (12)(13). Of note, GRS is positive, while GLS and GCS are negative. Since larger absolute values indicate greater myocardial deformation, all strain metrics are reported as absolute values for consistency and clarity.

Left atrial indexed volumes (LAVi) and derived indices of function were included. The indexed volumes were maximum (LAVi max), minimum (LAVi min), and pre-atrial contraction (LAVi preA). Phasic left atrial (LA) function indices derived from these volumes included total, passive, and active LA emptying fraction (LAEF), reflecting reservoir, conduit, and booster pump functions (14,15). The LA expansion index, which reflects the LV reservoir function, was also calculated using a formula described elsewhere (15–17).

Markers of arterial function, including total arterial compliance, aortic distensibility, and systemic vascular resistance, were analysed to estimate ventricular-arterial coupling and changes in HTN.

### Ascertainment of outcomes

Clinical outcomes occurring after the imaging visit (incident events) were identified using specific UK Biobank fields (Supplemental Table 3). Survival analyses involved censoring individuals based on the event date, date of death, or end of follow-up (30 November 2022), whichever came first. The clinical endpoints of interest included all-cause HF, all-cause mortality, AF, and atherosclerotic events. All-cause HF comprised HF, pulmonary oedema, or any cardiomyopathy as a possible cause (18). Atherosclerotic events included non-fatal MI, stroke, or peripheral artery disease. A composite of major adverse cardiovascular events (MACE) was also assessed, including HF, AF, atherosclerotic events, or cardiac death.

### Analysis workflow

The overall analysis comprised three main steps outlined below: data preparation, clustering, and *post hoc* statistical analyses.

#### Data preparation

From the 79 clinical features scrutinised as potential input for clustering (Supplemental Table 2), those with more than 20% missingness (microalbumin in urine) were excluded. As the cohort was mostly composed of Caucasian individuals (97%), ethnicity was also excluded as an input variable for clustering. This left 77 clinical features for clustering. Among these, the remaining missing values were imputed based on data type (mean/median for continuous data and most frequent value for categorical data). The cleaned dataset was then utilised as input for the clustering analysis.

#### Clustering analysis

Multiple clustering approaches were tested to determine the method that best balanced clustering performance, computational efficiency, and clinical interpretability. Further details on the clustering analysis are provided in the Supplemental Materials.

Briefly, the following approaches were explored: (1) K-Prototypes applied directly to the full dataset of 77 mixed-type variables, and to a reduced version (n = 65) after excluding highly correlated features based on a correlation matrix; (2) dimensionality reduction using Factorial Analysis of Mixed Data (FAMD), followed by clustering using K-Means; and (3) repeating the second approach on the reduced dataset (n = 65) after removing highly correlated variables. All approaches produced consistent results regarding the optimal number of clusters (two or three) across multiple evaluation metrics. However, the combination of FAMD for dimensionality reduction with K-Means clustering (approach 2) offered the best trade-off between model performance, computational efficiency, and clinical interpretability, and was therefore selected for the final analysis. The 40 FAMD-derived principal components, which explained over 80% of the total variance, were used as input for clustering. A three-cluster solution (*k* = 3) was selected based on the elbow method (which was consistent across all approaches), a Silhouette score similar to *k* = 2 (which showed the highest value), and the emergence of clinically meaningful, more granular phenotypic profiles.

Clustering stability was evaluated using bootstrapping and the Adjusted Rand Index (ARI), confirming robust and well-preserved cluster structures. To enhance interpretability and further confirm cluster assignments, we used Shapley Additive Explanations (SHAP) method, an explainable artificial intelligence technique that quantifies the contribution of individual features to model predictions. SHAP was applied to a supervised classifier trained on the 40 FAMD-derived principal components to predict the unsupervised cluster labels. This approach helped identify the components, and the underlying features they represent, that were most influential in driving cluster separation. The top 32 features contributing to cluster differentiation were then identified and ranked based on their contribution to the principal components (Table 1). These SHAP-ranked features were used to visualise and explain the main differences between clusters, improving clinical interpretability.

**Table 1.**
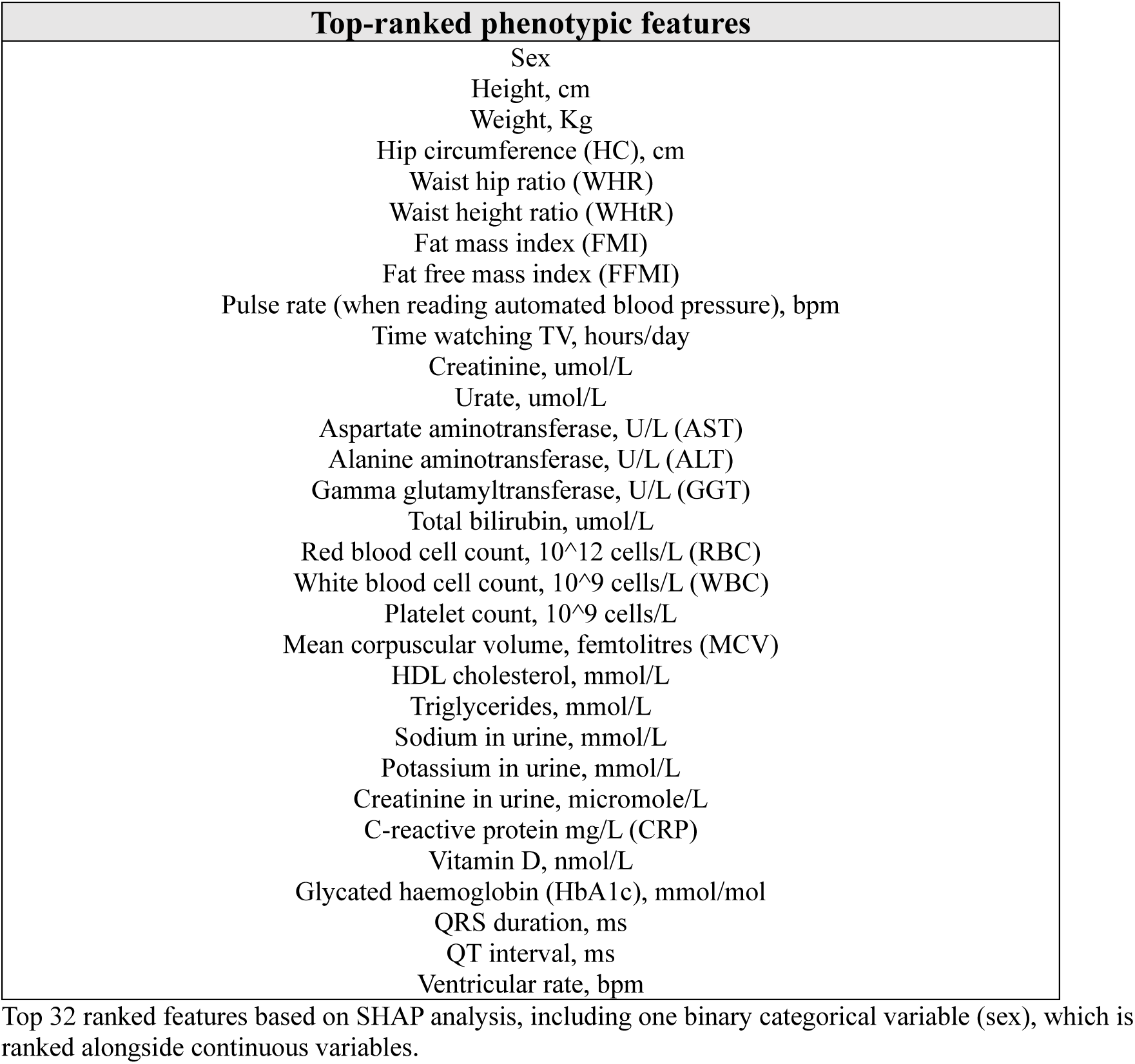
Top-ranked phenotypic features contributing to clustering.

| Top-ranked phenotypic features |
| --- |
| Sex |
| Height, cm |
| Weight, Kg |
| Hip circumference (HC), cm |
| Waist hip ratio (WHR) |
| Waist height ratio (WHtR) |
| Fat mass index (FMI) |
| Fat free mass index (FFMI) |
| Pulse rate (when reading automated blood pressure), bpm |
| Time watching TV, hours/day |
| Creatinine, umol/L |
| Urate, umol/L |
| Aspartate aminotransferase, U/L (AST) |
| Alanine aminotransferase, U/L (ALT) |
| Gamma glutamyltransferase, U/L (GGT) |
| Total bilirubin, umol/L |
| Red blood cell count, $10^{12}$ cells/L (RBC) |
| White blood cell count, $10^9$ cells/L (WBC) |
| Platelet count, $10^9$ cells/L |
| Mean corpuscular volume, femtolitres (MCV) |
| HDL cholesterol, mmol/L |
| Triglycerides, mmol/L |
| Sodium in urine, mmol/L |
| Potassium in urine, mmol/L |
| Creatinine in urine, micromole/L |
| C-reactive protein mg/L (CRP) |
| Vitamin D, nmol/L |
| Glycated haemoglobin (HbA1c), mmol/mol |
| QRS duration, ms |
| QT interval, ms |
| Ventricular rate, bpm |

Top 32 ranked features based on SHAP analysis, including one binary categorical variable (sex), which is ranked alongside continuous variables.

#### *Post hoc* analyses

Clustering was performed independently of clinical outcomes and CMR metrics, which were then used in *post hoc* analyses to assess the clinical relevance of the identified phenotypes and better characterise underlying risk profiles. Associations between cluster membership (categorical predictor; cluster 1 as reference) and clinical outcomes were assessed using Cox proportional hazards models, ensuring there were no violations. Both unadjusted and adjusted models were fitted, accounting for potential confounders, including hypercholesterolaemia, diabetes, and previous MI. Hazard ratios (HR) were estimated, and Kaplan-Meier survival analyses with log-rank tests were used to compare event-free survival across clusters.

The relationship between clusters and CMR metrics was investigated using multivariable linear regression models, adjusting for the same confounders used in the outcome analyses. To examine whether CMR metrics mediated the association between clusters and outcomes, mediation analyses were conducted using logistic regression models (PROCESS, R & SPSS) (20). In the model, cluster membership was the categorical predictor, the outcome was the binary output, and each CMR metric was tested separately as a continuous mediator. All models were adjusted for the same confounders used in previous analyses. Total, direct, and indirect effects were expressed as odds ratios (ORs) with 95% confidence intervals. The proportion mediated by each CMR metric was then calculated using a previously published formula (21).

As the HTN cohort included some individuals with early-stage cardiomyopathy (n=1,513, including ischaemic and non-ischaemic), we repeated the *post hoc* analyses excluding them to ensure the clustering results reflected hypertensive risk profiles rather than underlying cardiomyopathy.

#### Statistical analysis

Clusters were compared on clinical characteristics using chi-squared tests for categorical variables and ANOVA (or Kruskal-Wallis when applicable) for continuous variables.

Pairwise comparisons were performed with independent t-tests (for normally distributed data) or Wilcoxon rank-sum tests (for skewed data) for numeric variables, along with chi-squared tests for categorical variables. A two-sided p < 0.05 with Bonferroni correction for multiple testing, where appropriate, was deemed statistically significant for all analyses. All analyses were conducted using Python 3.8.10 (Python Software Foundation, Delaware USA) and Scikit-learn version 0.23.2 (19).

## Results

### Study population characteristics

The HTN cohort (n=14,840) was predominantly middle-aged (66.56±7.24 years), Caucasian (97%), and included 42% of females. Cardiovascular risk factors were prevalent, including hypercholesterolaemia (53%), smoking (17 %), and diabetes (12%). The prevalence of MI was approximately 11% (Table 2). Participants had borderline blood pressure values (systolic blood pressure [SBP], 147.1 ±18.3 mmHg; diastolic blood pressure [DBP], 81.5±10.3 mmHg). Over a median follow-up of 8.3 years, 591(3.98%) subjects developed HF, 778 (5.24%) experienced atherosclerotic events, 366 (2.46%) were diagnosed with AF, and 244 (1.64%) died. The overall incidence of MACE was 8.84% (n=1,312).

**Table 2.** Baseline clinical characteristics of the study cohort.

| Baseline characteristics | All cohort<br>(n = 14,840) |
| --- | --- |
| ***Demographics*** |  |
| Age, years | 66.56±7.24 |
| Female, n (%) | 6217(41.89) |
| Ethnicity, n (%) |  |
| *White* | 14368(96.82) |
| *Others* | 471(3.17) |
| ***Comorbidities, n (%)*** |  |
| Smoking history | 2586(17.42) |
| Previous myocardial infarction | 1608(10.83) |
| Stroke | 585(3.94) |
| Chronic obstructive pulmonary disease | 391(2.63) |
| Asthma | 2216(14.93) |
| Atrial fibrillation | 736(4.96) |
| Peripheral artery disease | 319(2.15) |
| Diabetes | 1790(12.06) |
| Hypercholesterolemia | 7898(53.22) |
| CKD | 657(4.43) |
| ***Physical measurements*** |  |
| Body mass index, Kg/m<sup>2</sup> | 27.48(25.03-30.46) |
| Waist hip ratio | 0.89±0.08 |
| Systolic blood pressure, mmHg | 147.08 ±18.3 |
| Diastolic blood pressure, mmHg | 81.51±10.28 |
| ***Laboratories*** |  |
| eGFR, mL/min/1.73m<sup>2</sup> | 89.98 ±12.81 |
| LDL cholesterol, mmol/L | 3.52 ±0.87 |
| Triglycerides, mmol/L | 1.60(1.14-2.27) |

Values are mean (±standard deviation) when continuous or number (percentage) when categorical. Data are presented as median (interquartile range) where absolute skew is ≥ 0.9. Define any acronyms used e.g. CKD etc..

### Identifying distinct clusters

Clustering stratified the hypertensive cohort into three phenotypically distinct groups, as determined by the elbow method and supported by Silhouette scores and clinical interpretability (see Supplemental Materials). Cluster stability was high (ARI = 0.80), indicating strong agreement across resamplings. The resulting groups showed significant differences across a wide range of clinical, behavioural, laboratory, and ECG characteristics (Table 3; Supplemental Tables 4,5).

**Table 3.** Baseline clinical characteristics stratified by clusters.

|  | Cluster 1<br>(n=4300) | Cluster 2<br>(n=8339) | Cluster 3 (n=2201) | p Value | Cluster 1 vs Cluster<br>2 | Cluster 1 vs Cluster<br>3 | Cluster 2 vs Cluster<br>3 |
| --- | --- | --- | --- | --- | --- | --- | --- |
| ***Socio-demographics*** |  |  |  |  |  |  |  |
| Age, years | 66.75±7.20 | 66.84±7.24 | 65.14±7.20 | <0.001 | 1 | <0.001 | <0.001 |
| Female, n (%) | 4117(95.74) | 8(0.10) | 2092(95.05) | <0.001 | <0.001 | 0.66 | <0.001 |
| Townsend deprivation index | -2.6(-3.8,-0.4) | -2.7(-3.9,-0.5) | -2.1(-3.6,-0.4) | <0.001 | 1 | <0.001 | <0.001 |
| Educational level, n (%) |  |  |  | <0.001 | <0.001 | <0.001 | <0.001 |
| High | 2225(51.74) | 4459(53.47) | 988(44.89) |  |  |  |  |
| Intermediate | 1502(34.93) | 2465(29.56) | 843(38.30) |  |  |  |  |
| Low | 573(13.32) | 1415(19.96) | 370(16.81) |  |  |  |  |
| ***Comorbidities, n (%)*** |  |  |  |  |  |  |  |
| Previous myocardial infarction | 229(5.32) | 1228(14.72) | 151(6.86) | <0.001 | <0.001 | <0.001 | <0.001 |
| Stroke | 114(2.65) | 393(4.71) | 78(3.54) | <0.001 | <0.001 | 0.16 | 0.06 |
| Chronic obstructive pulmonary disease | 70(1.63) | 239(2.87) | 82(3.72) | <0.001 | <0.001 | <0.001 | 0.13 |
| Asthma | 6.26(14.56) | 1161(13.92) | 429(19.49) | <0.001 | 1.03 | <0.001 | <0.001 |
| Atrial fibrillation | 130(3.02) | 518(6.21) | 88(4) | <0.001 | <0.001 | 0.14 | <0.001 |
| Peripheral artery disease | 101(2.35) | 182(2.18) | 36(1.63) | 0.49 | 1.78 | 0.21 | 0.39 |
| Diabetes | 198(4.60) | 1117(13.39) | 475(21.58) | <0.001 | <0.001 | <0.001 | <0.001 |
| Hypercholesterolemia | 1698(39.48) | 5103(61.19) | 1097(49.84) | <0.001 | <0.001 | <0.001 | <0.001 |
| Chronic kidney disease | 164(3.81) | 363(4.35) | 130(5.91) | 0.001 | 0.49 | <0.001 | 0.007 |
| ***Physical measurements*** |  |  |  |  |  |  |  |
| Waist hip ratio | 0.80±0.06 | 0.94±0.06 | 0.87±0.07 | <0.001 | <0.001 | <0.001 | <0.001 |
| Fat mass index, kg/m<sup>2</sup> | 8.66(7.07-10.21) | 7.23(5.88-8.75) | 14.21(12.6-16.48) | <0.001 | <0.001 | <0.001 | <0.001 |
| Fat free mass index, kg/m<sup>2</sup> | 16.49±1.23 | 20.75±1.76 | 18.92±2.04 | <0.001 | <0.001 | <0.001 | <0.001 |

|  | **Cluster 1**<br>**(n=4300)** | **Cluster 2**<br>**(n=8339)** | **Cluster 3 (n=2201)** | **p Value** | **Cluster 1 vs Cluster**<br>**2** | **Cluster 1 vs Cluster**<br>**3** | **Cluster 2 vs Cluster**<br>**3** |
| --- | --- | --- | --- | --- | --- | --- | --- |
| DBP, mmHg | 80.74±9.39 | 82.05±9.32 | 80.97±9.03 | <0.001 | <0.001 | 1 | <0.001 |
| SBP, mmHg | 147.32±17.72 | 147.41±15.98 | 145.37±16.68 | <0.001 | 0.13 | <0.001 | <0.001 |
| Pulse rate, bpm | 70.53±10.21 | 68.35±11.31 | 73.3±11.34 | <0.001 | <0.001 | <0.001 | <0.001 |

Categorical values are presented as count (percentage); continuous values are presented as mean ( ± standard deviation) and as median (interquartile range) where the absolute skew is $\geq 0.9$ . The P Value indicates comparisons of variables across clusters, and bold values indicate statistical significance ( $p < 0.05$ ). Pairwise comparisons were calculated with Bonferroni (for multiple corrections). P value for continuous was calculated using ANOVA (or t-test for pairwise comparisons) if normally distributed, and with Kruskal-Wallis (or x for pairwise comparisons) if not normally distributed. Categorical variables were compared using a chi-squared test for independence. *DBP* = diastolic blood pressure, *SBP* = systolic blood pressure.

Cluster 1 (n=4,300) was predominantly female (95.7%) with leaner body composition, lower diabetes (4.6%) and hypercholesterolaemia (39.5%) rates, and moderate smoking history (35.4%). This group exhibited better lifestyle habits, a more favourable metabolic profile (e.g., higher HDL and lower triglycerides), and decreased systemic inflammation (C-reactive protein). They had distinct ECG features, such as a shorter QRS duration than cluster 2 and a lower resting heart rate than cluster 3.

Cluster 2 (n=8,339) was mainly male (99.9%) and exhibited significant CV disease burden, including MI (14.7%) and AF (6.2%). This group had a high waist-to-hip ratio but a lower fat mass index than cluster 3. They also had the lowest LDL and HDL cholesterol levels and signs of subclinical kidney dysfunction. Lifestyle habits included high smoking rates (49.4%) and alcohol and processed food consumption. Their ECG profile showed the longest QRS duration (90 ms) and the lowest resting pulse rate.

Cluster 3 (n=2,201) was predominantly female, like cluster 1, but younger (65.14±7.20 years), with the highest central adiposity, and greatest prevalence of diabetes (21.6%). They had lower physical activity, greater socioeconomic deprivation, and the highest systemic inflammatory markers. This group showed higher resting heart rate (64 bpm) and shorter QRS duration (86 ms) than cluster 2. With regard to the blood pressure values, cluster 3 showed the lowest SBP (145.37 mmHg), while cluster 2 displayed the highest DBP (82.05 mmHg) among the groups.

The top 32 SHAP features identified for effective clustering summarised diverse clinical phenotypes (Figure 1). They included body composition, metabolic markers, cardiovascular characteristics, and lifestyle habits, reflecting each cluster’s risk profile. This confirms SHAP as a valuable tool for capturing core characteristics and enhancing interpretability while facilitating meaningful comparisons between clusters.

**Figure 1.**
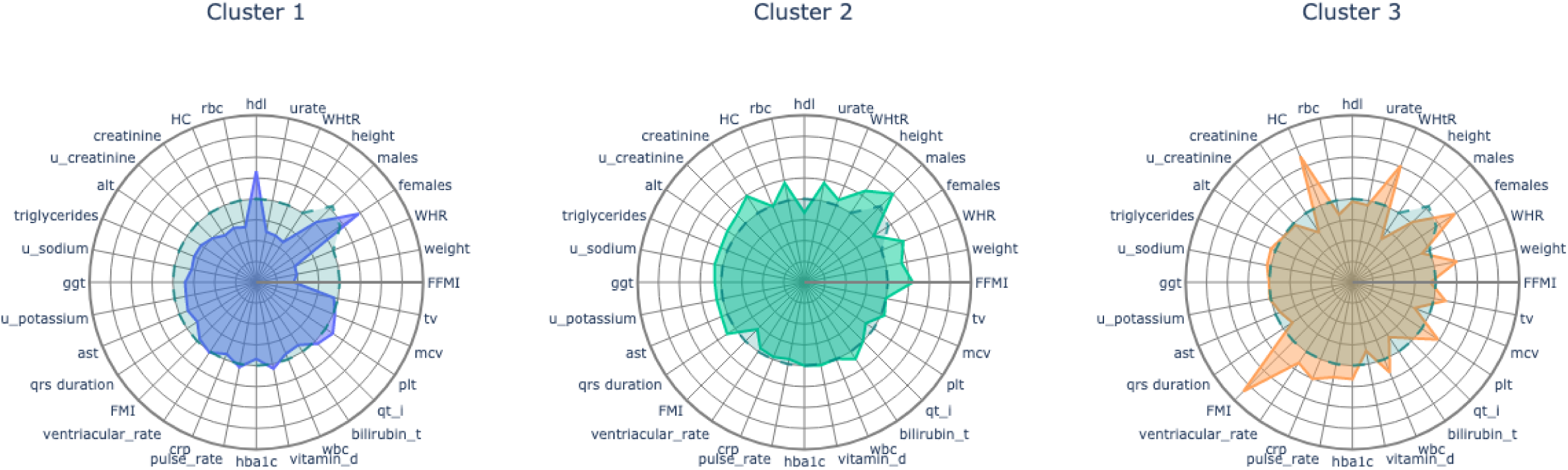
Radar charts showing the top 32 features contributing to clustering. Radar plots summarising the top 32 (scaled) features contributing to clustering obtained from SHAP compared to their average value for the entire cohort (indicated by the dashed blue line). Abbreviations are explained in Table 1.

### Cluster associations with clinical outcomes

Kaplan-Meier analyses revealed significantly different survival trajectories across clusters for all adverse events (Figure 2, multivariate log-rank test p < 0.005). Cluster 1 exhibited the lowest risk, cluster 2 the highest, and cluster 3 an intermediate risk profile.

**Figure 2.**
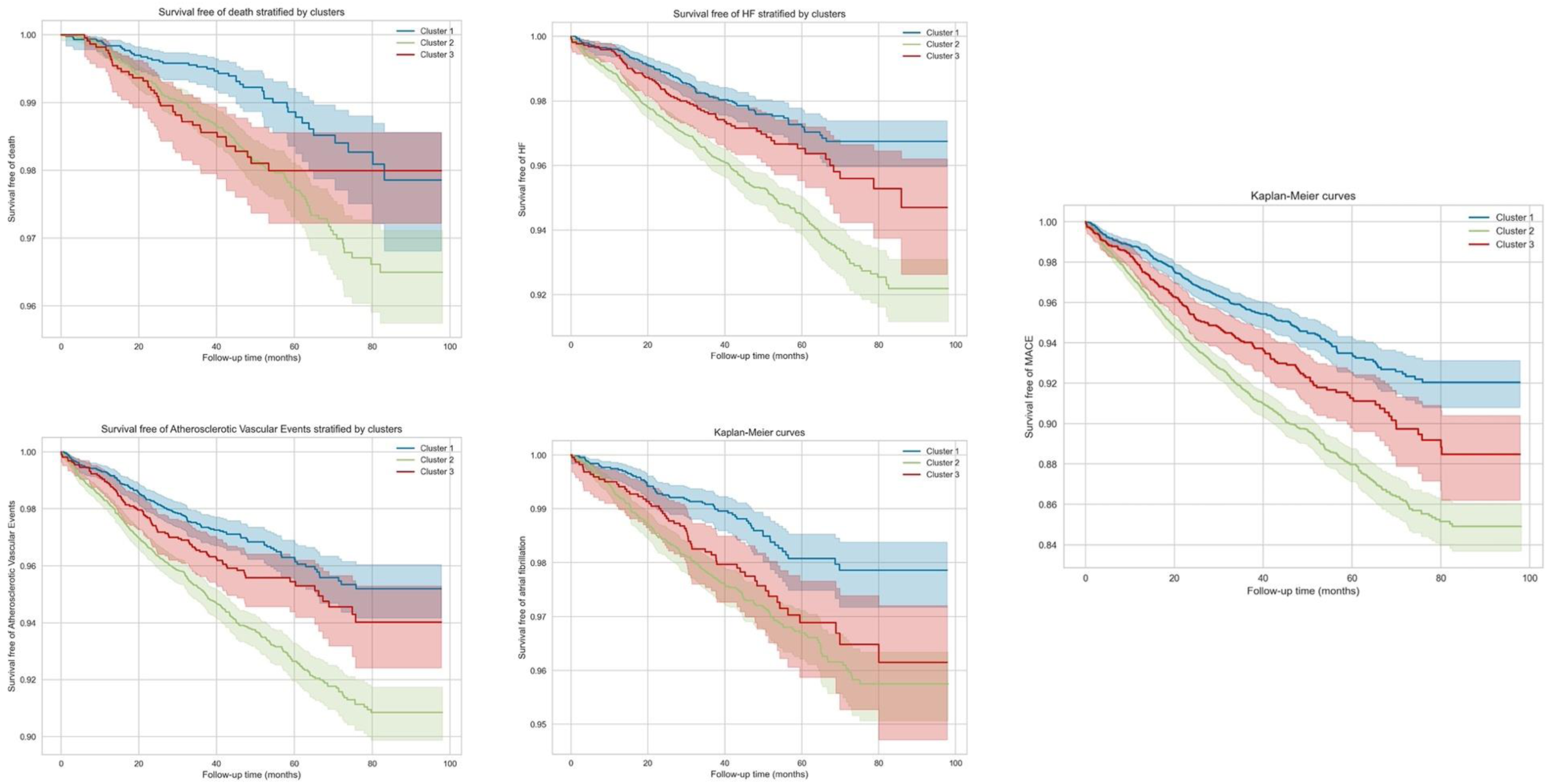
Kaplan-Meier survival analysis for incident events stratified by clusters.

Cox proportional hazard models confirmed this finding (Table 4). Cluster 2 had the greatest risk for all adverse events compared to cluster 1 (the lowest risk). The highest HRs were observed for HF (HR: 1.80 [95% CI: 1.45–2.25], p < 0.005) and AF (HR: 1.85 [95% CI: 1.39–2.46], p < 0.005). Cluster 3 showed moderate risk for AF (HR: 1.61 [95% CI: 1.12–2.32], p < 0.05) and MACE (HR: 1.30 [95% CI: 1.07–1.58], p < 0.05) after adjusting for cardiovascular risk factors.

**Table 4.**
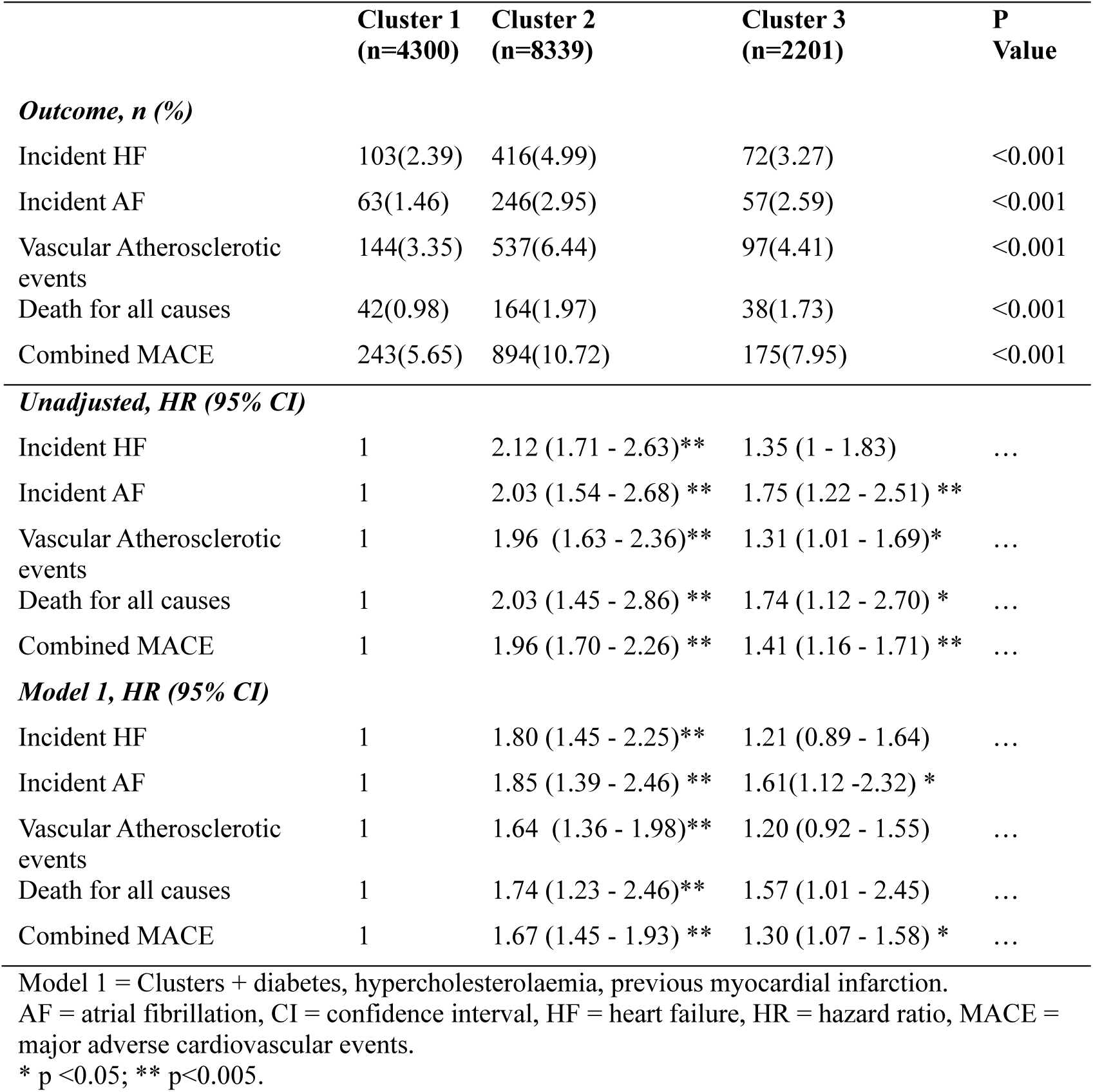
Association of clusters with adverse outcomes on Cox proportional **hazards analysis.**

|  | **Cluster 1**<br>**(n=4300)** | **Cluster 2**<br>**(n=8339)** | **Cluster 3**<br>**(n=2201)** | **P**<br>**Value** |
| --- | --- | --- | --- | --- |
| ***Outcome, n (%)*** |  |  |  |  |
| Incident HF | 103(2.39) | 416(4.99) | 72(3.27) | <0.001 |
| Incident AF | 63(1.46) | 246(2.95) | 57(2.59) | <0.001 |
| Vascular Atherosclerotic events | 144(3.35) | 537(6.44) | 97(4.41) | <0.001 |
| Death for all causes | 42(0.98) | 164(1.97) | 38(1.73) | <0.001 |
| Combined MACE | 243(5.65) | 894(10.72) | 175(7.95) | <0.001 |
| ***Unadjusted, HR (95% CI)*** |  |  |  |  |
| Incident HF | 1 | 2.12 (1.71 - 2.63)** | 1.35 (1 - 1.83) | ... |
| Incident AF | 1 | 2.03 (1.54 - 2.68) ** | 1.75 (1.22 - 2.51) ** | ... |
| Vascular Atherosclerotic events | 1 | 1.96 (1.63 - 2.36)** | 1.31 (1.01 - 1.69)* | ... |
| Death for all causes | 1 | 2.03 (1.45 - 2.86) ** | 1.74 (1.12 - 2.70) * | ... |
| Combined MACE | 1 | 1.96 (1.70 - 2.26) ** | 1.41 (1.16 - 1.71) ** | ... |
| ***Model 1, HR (95% CI)*** |  |  |  |  |
| Incident HF | 1 | 1.80 (1.45 - 2.25)** | 1.21 (0.89 - 1.64) | ... |
| Incident AF | 1 | 1.85 (1.39 - 2.46) ** | 1.61(1.12 -2.32) * | ... |
| Vascular Atherosclerotic events | 1 | 1.64 (1.36 - 1.98)** | 1.20 (0.92 - 1.55) | ... |
| Death for all causes | 1 | 1.74 (1.23 - 2.46)** | 1.57 (1.01 - 2.45) | ... |
| Combined MACE | 1 | 1.67 (1.45 - 1.93) ** | 1.30 (1.07 - 1.58) * | ... |

Model 1 = Clusters + diabetes, hypercholesterolaemia, previous myocardial infarction.
AF = atrial fibrillation, CI = confidence interval, HF = heart failure, HR = hazard ratio, MACE = major adverse cardiovascular events.
\* p <0.05; \*\* p<0.005.

### Associations between clusters and CMR metrics

CMR analysis revealed significant differences across the clusters (Table 5 and Figure 3), with cluster 1 showing the most favourable CV imaging features. Cluster 2 was associated with the largest biventricular volumes, greatest cardiac remodelling (highest M/V ratio), and LV hypertrophy. It also had the poorest LA function (primarily passive LAEF and LA expansion index) and LV mechanics (GLS, GRS, GCS), especially the LV and RV longitudinal strains. Despite minimal arterial stiffening (highest total arterial compliance and aortic distensibility), it showed the greatest systemic vascular resistance. Cluster 3 had smaller biventricular volumes than the reference groups, milder LV remodelling, and LV hypertrophy compared to cluster 2, along with less reduction in LV mechanics (GCS and GRS) and better RV strains. They also showed a milder reduction in LA functions, including the LA booster pump function.

**Table 5.**
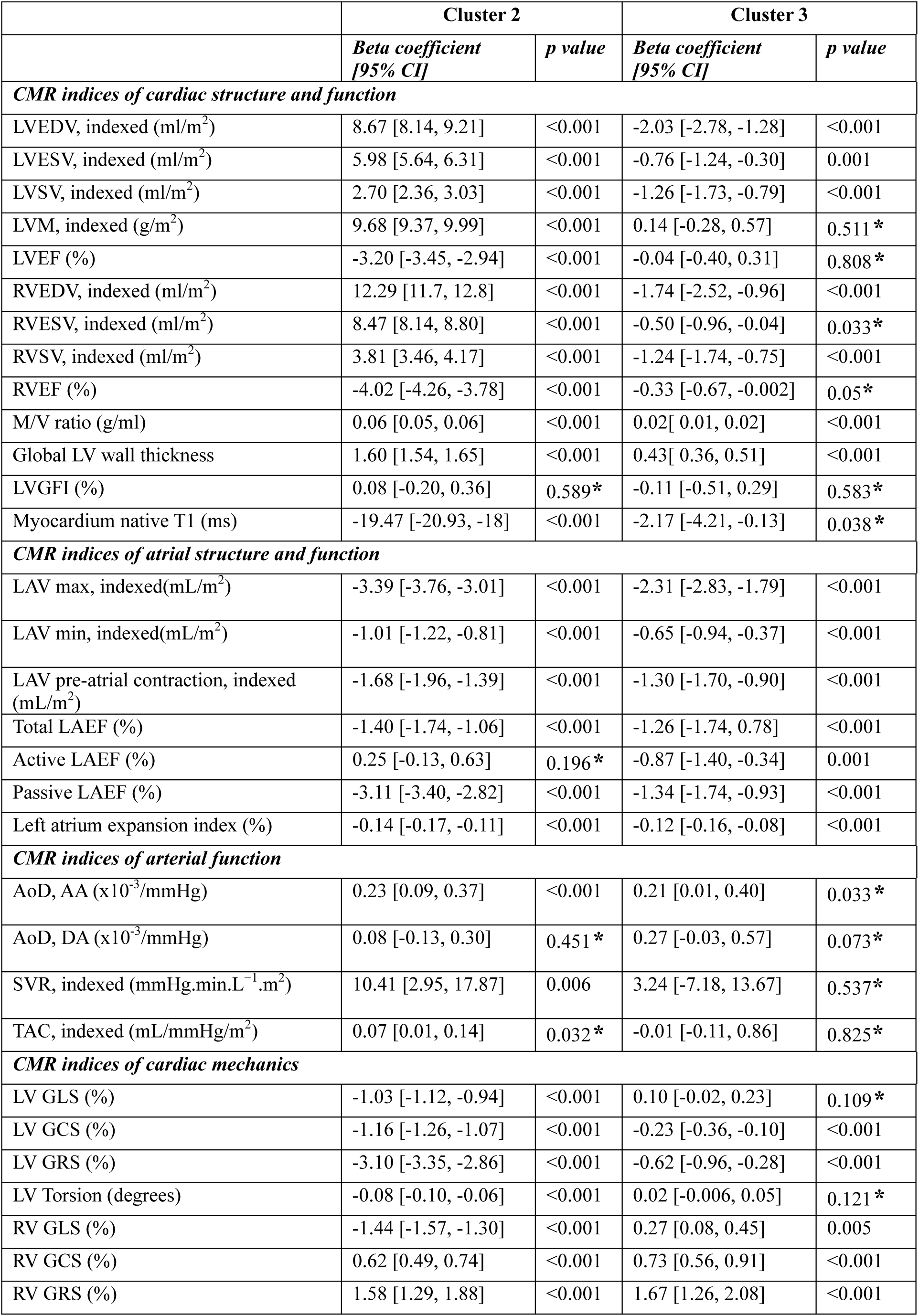
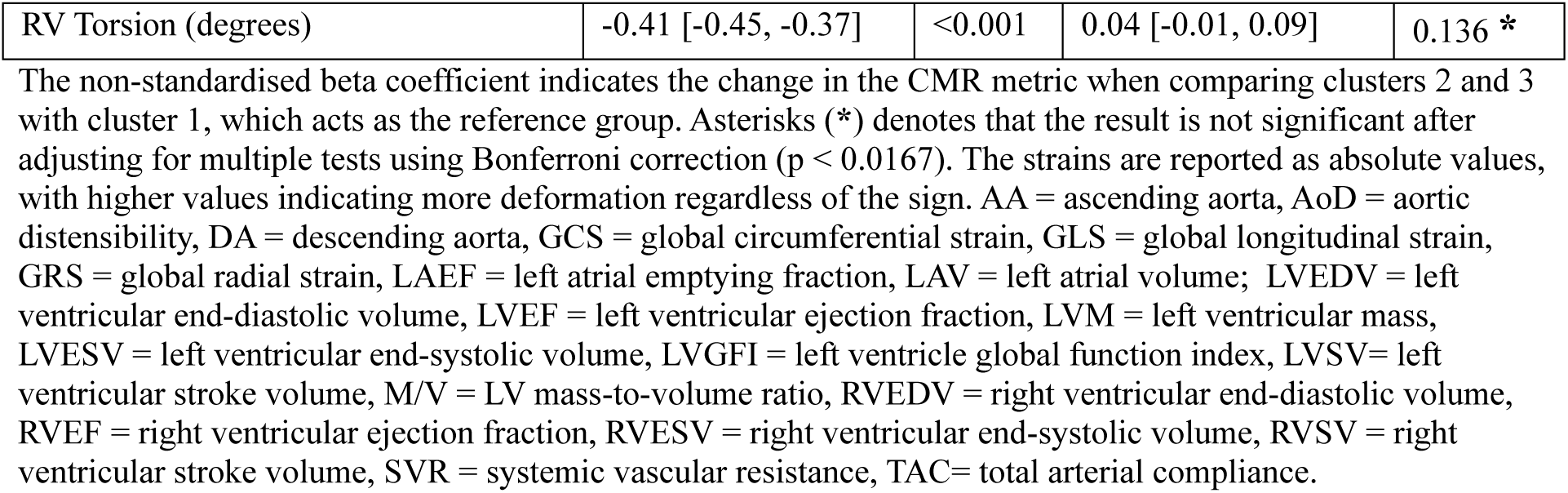
Associations of CMR metrics with clustering.

**Figure 3.**
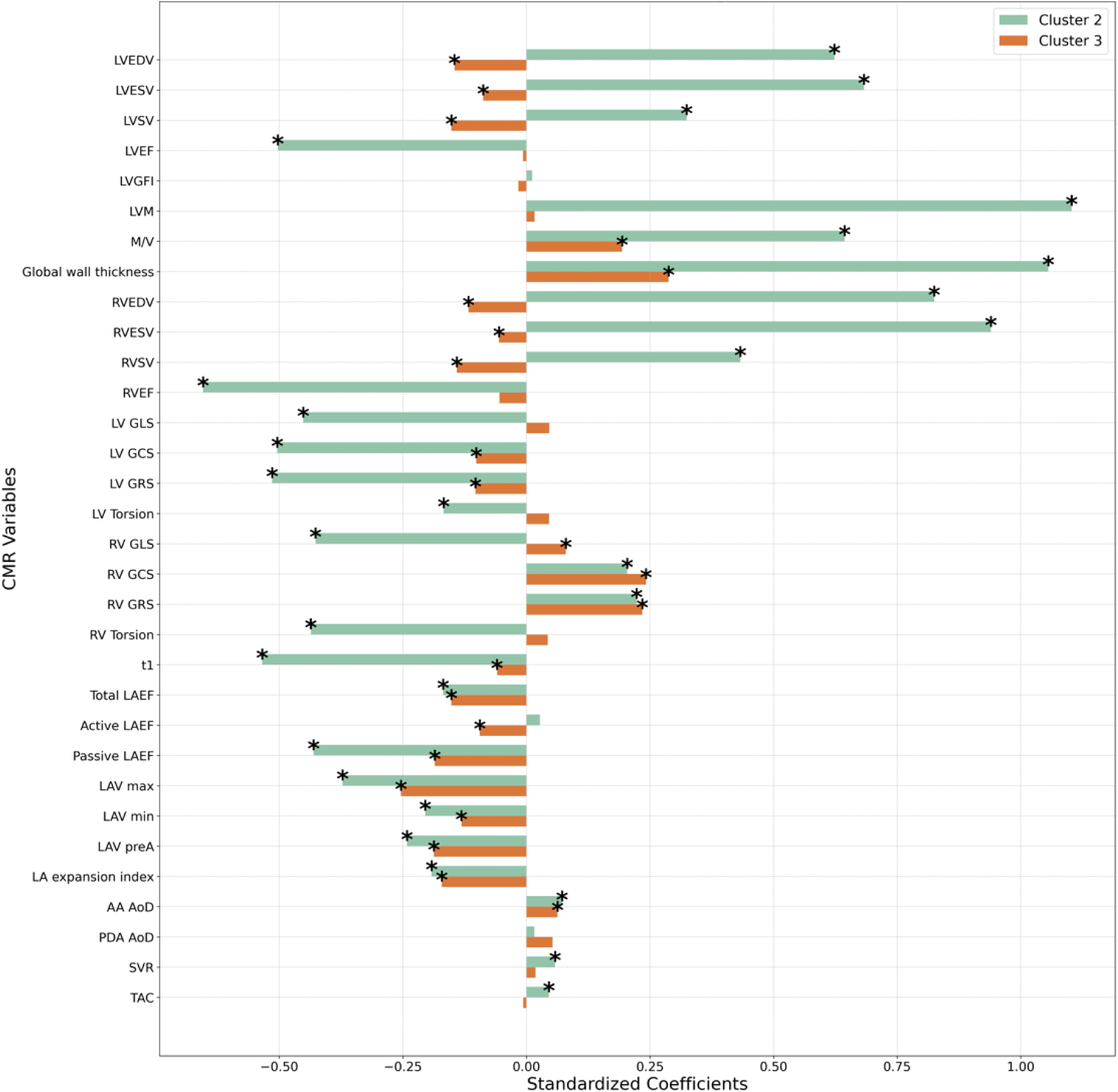
Associations between CMR metrics and clustering. This figure illustrates the associations between CMR-derived metrics and clustering, comparing cluster 2 and cluster 3 to the lowest-risk reference group (Cluster 1). The bars represent standardised beta coefficients from regression models, indicating the magnitude and direction of changes in each CMR metric when comparing clusters 2 and 3 to cluster 1 Positive coefficients suggest higher values of the respective CMR metric in clusters 2 or 3 compared to cluster 1, while negative coefficients indicate lower values relative to the reference group. Only significant associations after Bonferroni correction for multiple testing are displayed. Asterisks (*) indicate statistically significant associations per each group. Abbreviations are explained in Table 5.

### Mediation role of CMR metrics in the association between cluster and outcomes

In cluster 2, the highest proportion of MACE risk (Figure 4) was mediated (indirect effect) through LV hypertrophy (LV mass, 67%), geometric remodelling (M/V ratio, 20%), and LV volumes, alongside a reduction in LV ejection fraction (25%), LV mechanics, particularly GLS (33%), and alterations in passive LAEF (25%). In cluster 3, the indirect effect of CMR changes on MACE was considerably smaller, with the highest proportion mediated by LV maximum wall thickness (26%) rather than LV mass, and all indices of LA function, including LA expansion index (19%), total (17%), passive (15%), and active (9%) LAEF.

**Figure 4.**
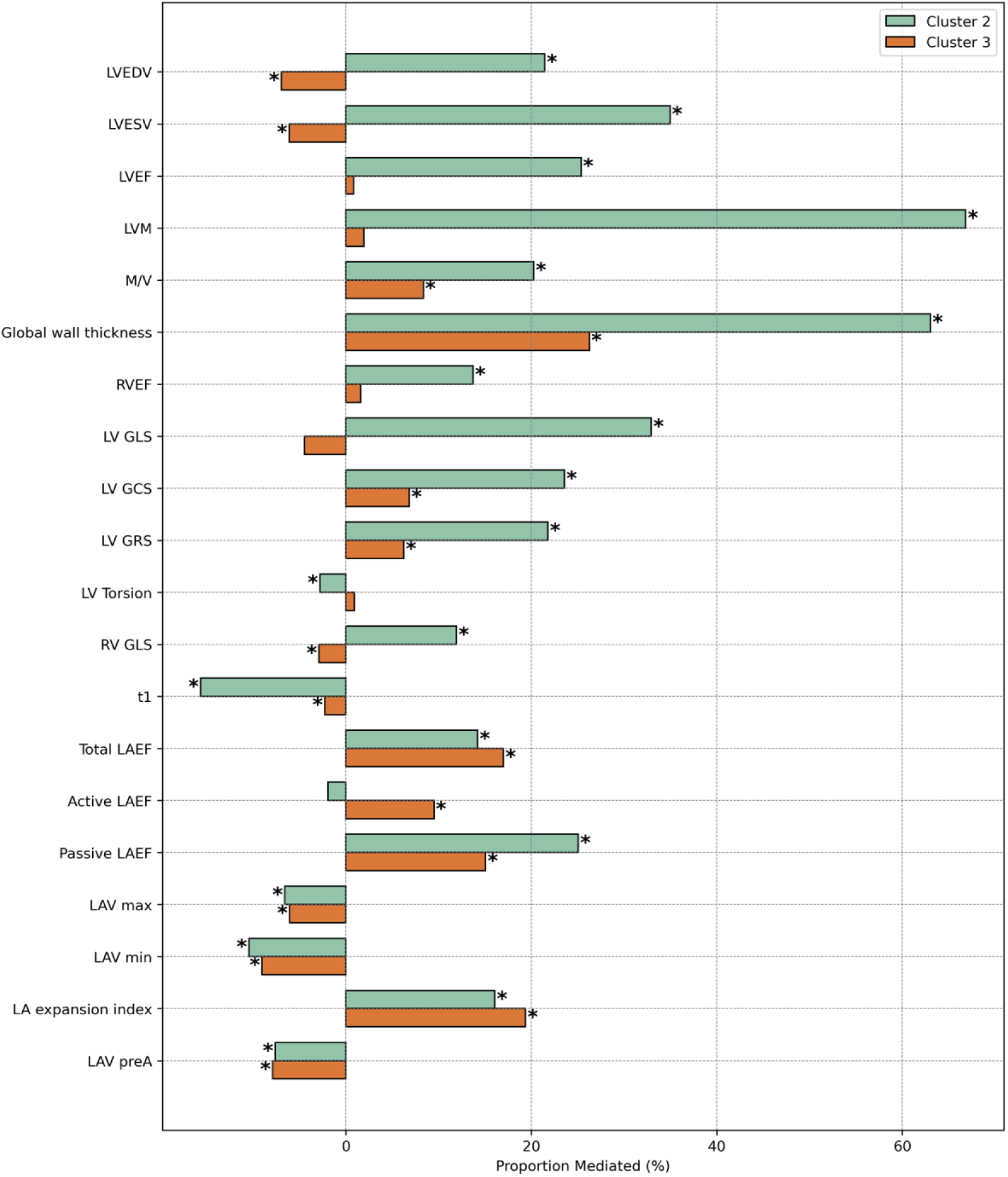
Proportion of MACE risk mediated by CMR features across clusters. The figure shows the proportion of the total effect of clustering on the outcome mediated through CMR metrics, expressed as a percentage. Positive values indicate mediation, where the CMR variable explains part of the association between clustering and the outcome. Negative values suggest a suppressor effect, strengthening the direct association. Cluster 2 (higher-risk) and cluster 3 (intermediate-risk) are compared to cluster 1 (lowest-risk reference). Asterisks (*) indicate statistically significant mediation effects.

Mediation patterns in HF (Figure 5) and atherosclerotic events (Figure 6) resembled those seen in MACE in both clusters; however, CMR metrics had a more pronounced mediation effect in HF, and especially in cluster 2, where LVH and cardiac remodelling were key contributors. In associations with AF (Figure 7), patterns mirrored MACE in both clusters, but indices of LA function became more prominent.

**Figure 5.**
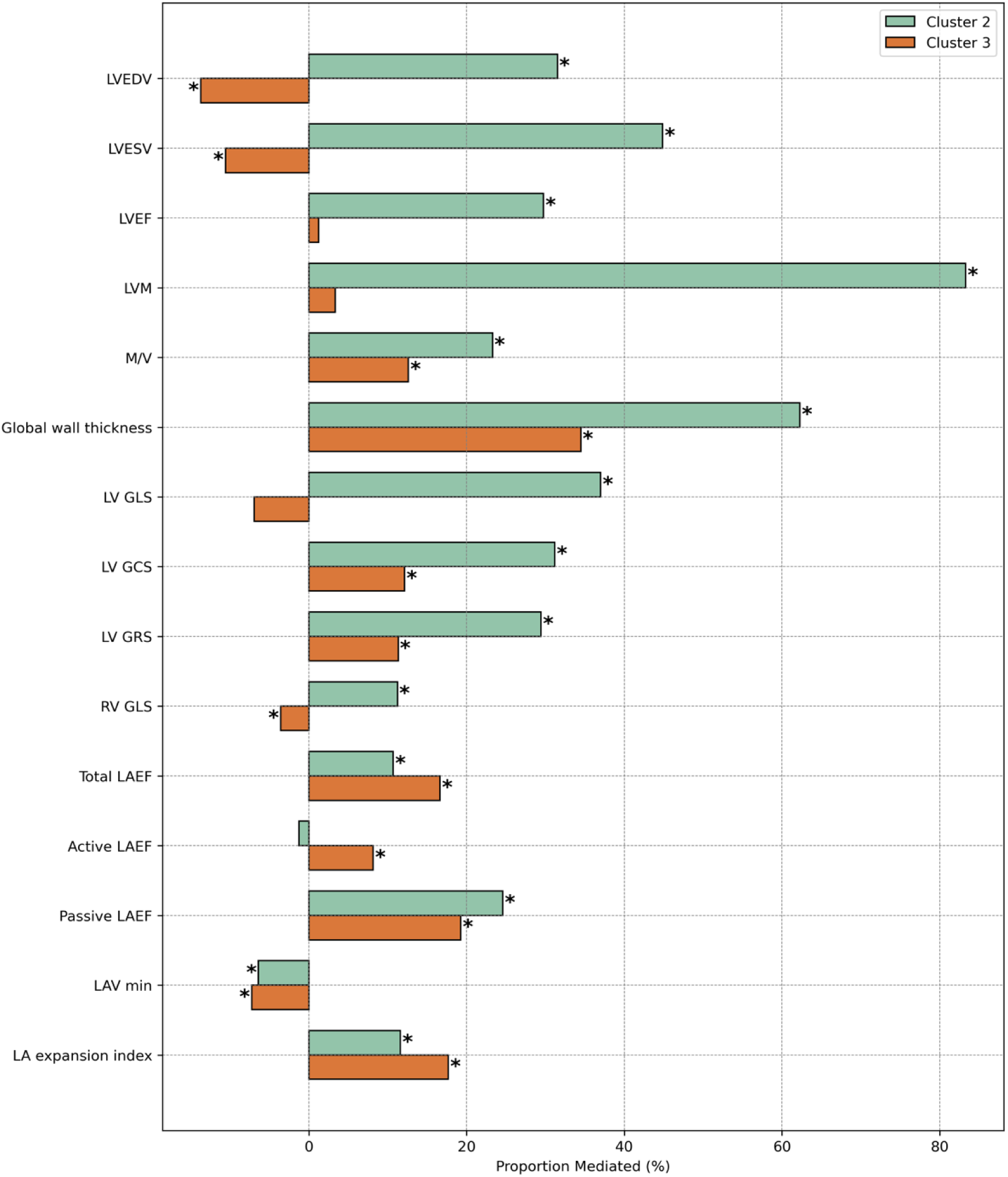
Proportion of HF risk mediated by CMR features across clusters. See Figure 4 for a detailed explanation of the mediation analysis and interpretation of the results.

**Figure 6.**
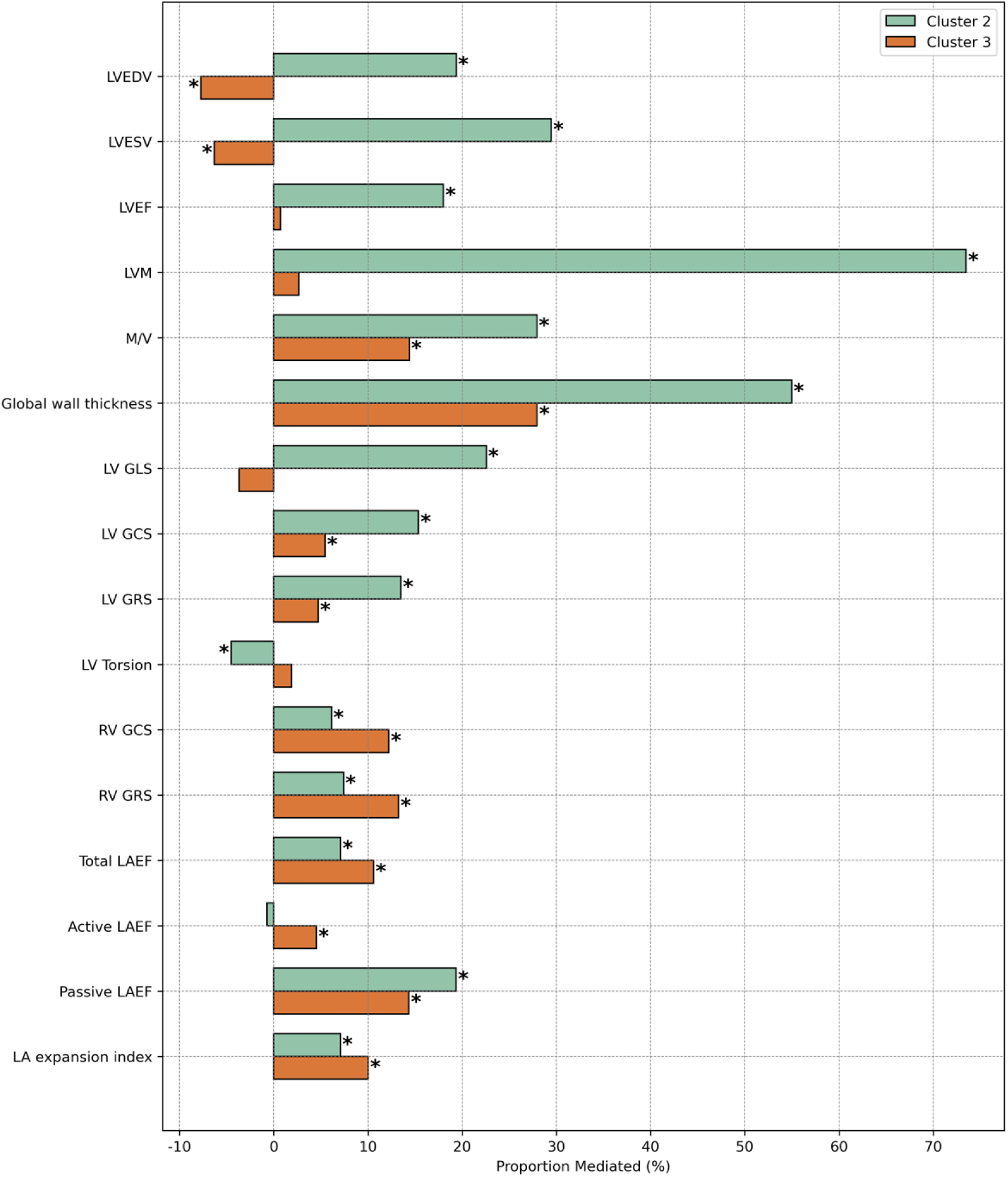
Proportion of the risk of incident atherosclerotic events mediated by CMR features across clusters. See Figure 4 for a detailed explanation of the mediation analysis and interpretation of the results.

**Figure 7.**
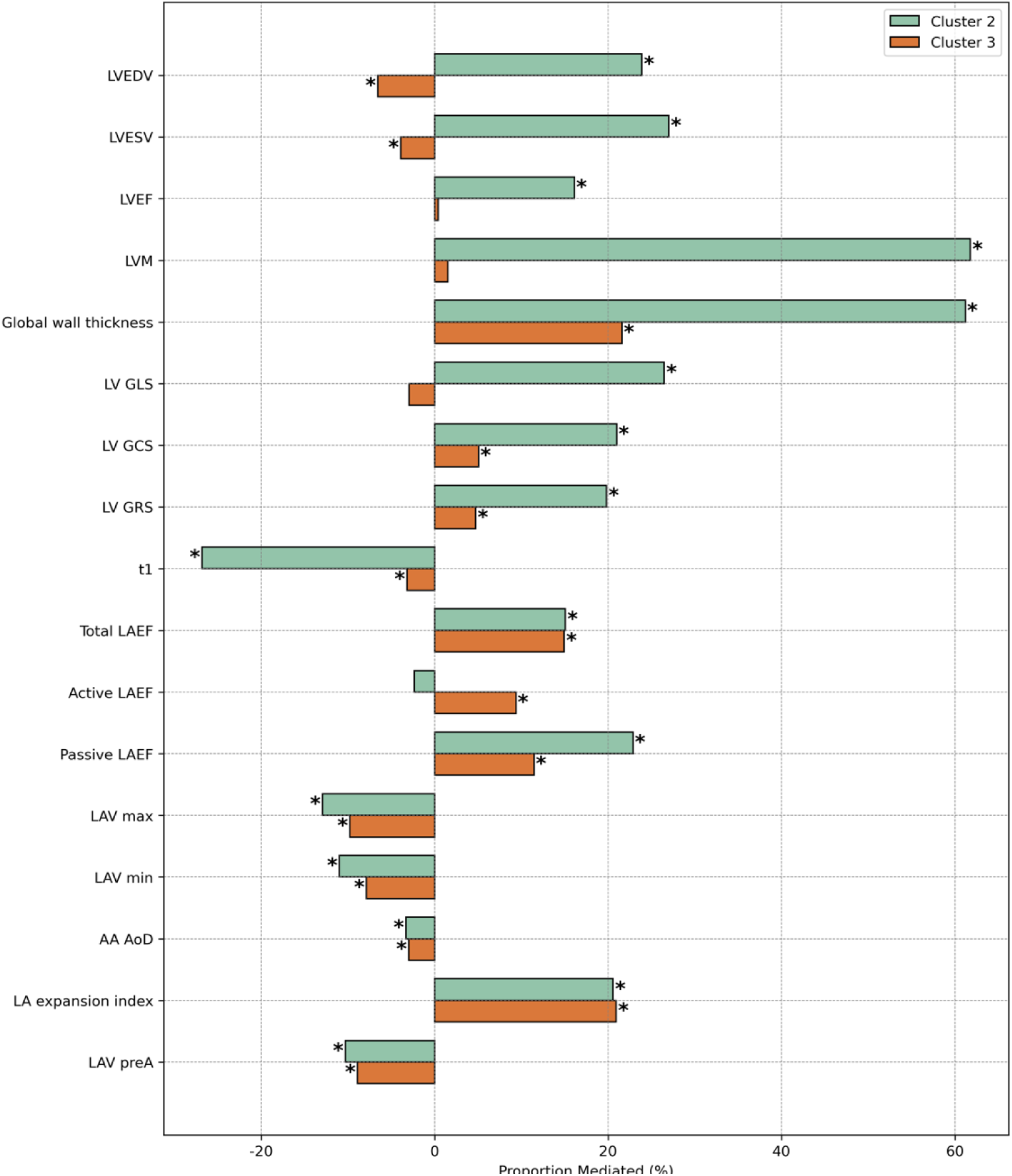
Proportion of AF risk mediated by CMR features across clusters. See Figure 4 for a detailed explanation of the mediation analysis and interpretation of the results.

The contributing role of CMR metrics to death risk was less pronounced (Figure 8). In Cluster 2, the impact of LA function indices (the greatest was the LA expansion index, 66%) and volumes outweighed that of LV hypertrophy and LV function indices, indicating that atrial dysfunction is a major contributor to mortality in this cluster. Conversely, changes in RV (−20%) and LV (−15%) stroke volumes, and T1 (−19%) served as suppressors. In cluster 3, the mediation patterns resembled those of MACE, but the indirect effects were minimal, representing the smallest proportions across all outcomes.

**Figure 8.**
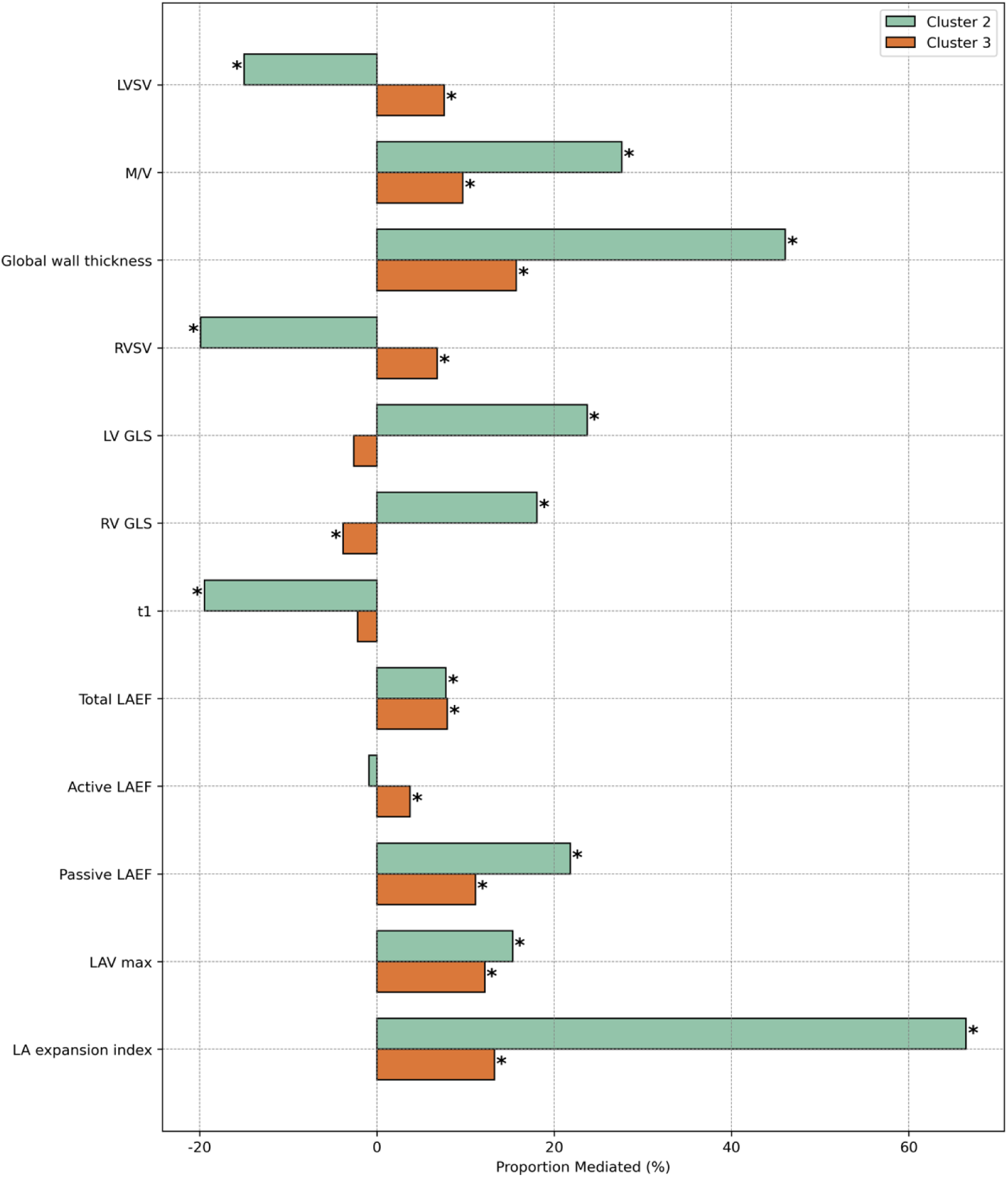
Proportion of death risk mediated by CMR features across clusters. See Figure 4 for a detailed explanation of the mediation analysis and interpretation of the results.

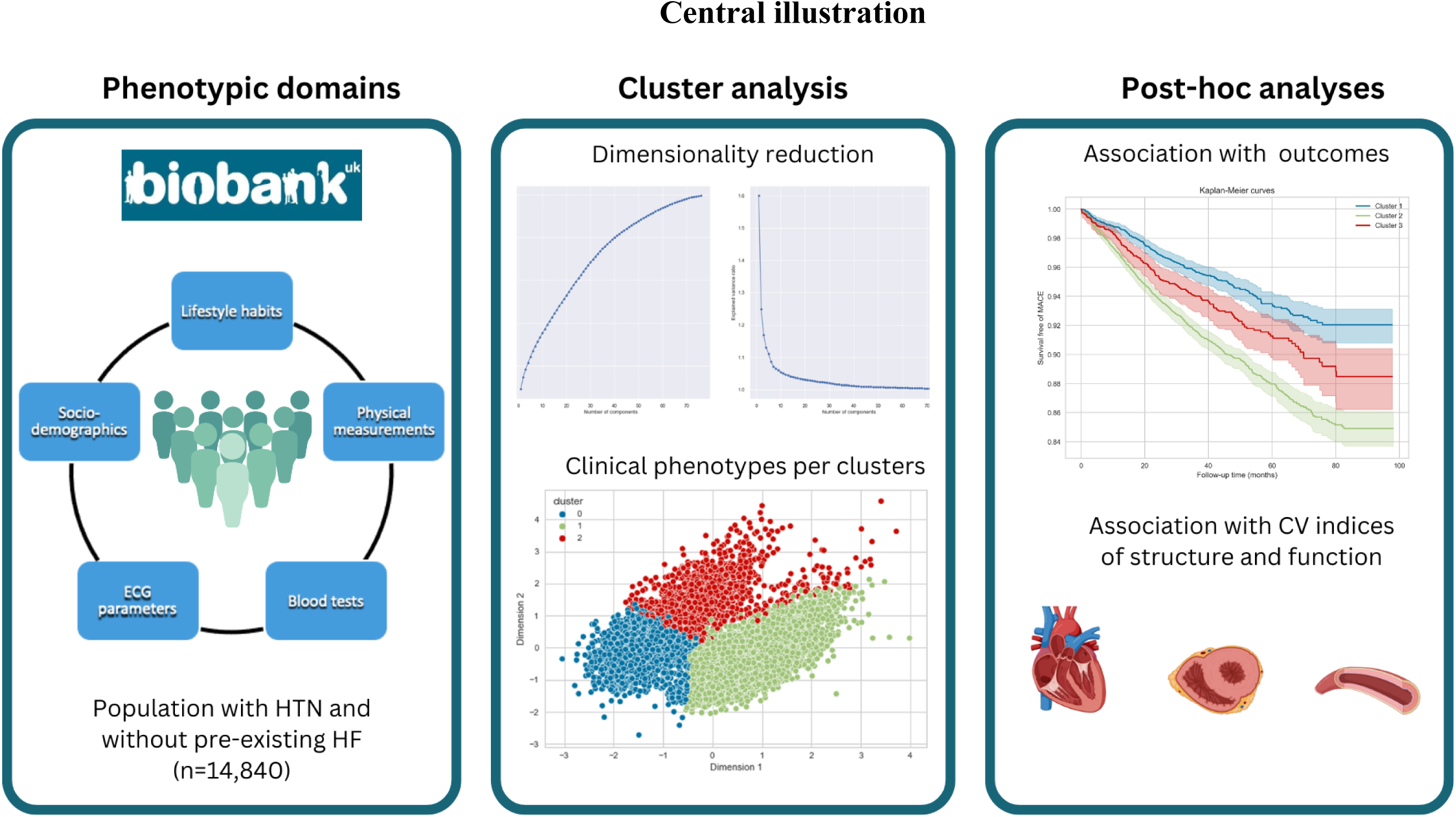
Central illustration. This illustration summarises the study, which applied clustering analysis to clinical and physiological data from hypertensive UK Biobank participants. Clustering identified three distinct phenotypes, shown in the middle panel. Post hoc analyses (right panel) validated these clusters by examining their associations with cardiovascular structure, function, and outcomes. The findings reveal distinct risk profiles and remodelling patterns, highlighting different mechanisms driving cardiovascular risk.

### Sensitivity analyses

There were 1,513 hypertensive participants with also cardiomyopathies, distributed as follows: cluster 1, 4.9%; cluster 2, 13.9%; and cluster 3, 6.54%. After removing these cases (n= 13,327), cluster 2 remained the highest-risk group for all adverse events. However, the greatest risks shifted from HF and AF to AF and all-cause mortality (HR: 1.97 [95% CI: 1.46-2.66] for AF, HR: 1.83 [95% CI: 1.26-2.64] for death; p <0.005). In Cluster 3, the AF risk remained unchanged (HR: 1.62 [95% CI: 1.10-2.29], p <0.005), while MACE risk was attenuated, and all-cause mortality became significant (HR: 1.67 [95% CI: 1.04-2.68], p <0.005) (Supplemental Table 6).

Associations of clusters with CMR metrics (n=10,873) and their outcome roles were consistent with previous analyses (Supplemental Figure 1). However, in cluster 3, LV GLS and the LA expansion index influenced all event risks more than before (Supplemental Figure 2). Notably, LA function and cardiac mechanics were the only significant mediators in both clusters for all-cause mortality. (Supplemental Figure 3). This suggests that including individuals with cardiomyopathies in cluster 3 masked the influence of atrial function and LV GLS on event risk. In cluster 2, their presence led to a slight overestimation of LV geometric remodelling as a mediator of mortality without significantly altering mediation effects for other outcomes.

## Discussion

In this large UK Biobank cohort, unsupervised clustering of clinically available data identified three HTN phenotypes with distinct risk profiles, CV imaging features, and outcomes despite modest blood pressure differences (Central illustration). The CV changes played varying roles in mediating risk, underscoring heterogeneity among hypertensive individuals and the potential of clustering for refining risk stratification.

### Three distinct hypertensive phenotypes

Cluster 1 represented the most favourable HTN phenotype, consisting mainly of females with healthier metabolisms and lifestyles. Despite some adverse vascular changes, like the lowest aortic compliance reflecting sex differences, they exhibited minimal cardiac remodelling and maintained good cardiac functions and mechanics (22). This milder HTN profile resulted in better clinical outcomes than other clusters, categorising this group as low risk.

Cluster 2, mainly male, exhibited an advanced atherosclerotic profile, marked by high MI prevalence, low HDL levels, and significant target organ damage (e.g. poor kidney function). Interestingly, LDL and total cholesterol were low, likely due to secondary prevention in severe atherosclerosis. This phenotype showed the most severe cardiac remodelling with extensive LV hypertrophy, larger ventricular volumes, and impaired LV mechanics (yet preserved LV ejection fraction), along with notable LA dysfunction. Although SBP was similar to cluster 1, DBP and SVR were the highest across the groups. The similarity in SBP, despite the higher risk profile of cluster 2, may reflect the impact of hypertensive therapy.

However, the elevated DBP and systemic vascular resistance, together with more advanced structural and functional abnormalities, likely indicate more severe vascular disease in cluster 2 due to combined LV hypertrophy and atherosclerosis, contributing to macro- and microvascular dysfunction that exacerbates LV dysfunction and increases the risk of adverse CV events (12,23). Hence, cluster 2 faced the greatest risk of adverse outcomes, particularly HF and AF. These findings support the relationship between LV hypertrophy, atherosclerosis, and adverse outcomes, especially in middle-aged males (24,25).

Cluster 3, which is predominantly female, exhibited key features of metabolic syndrome (MetS), such as central adiposity, poor glucose control, and systemic inflammation, alongside significant sociocultural challenges. While LV hypertrophy and atrial dysfunction were milder than in cluster 2, LA impairment was more extensive, affecting all functional phases. This group had the lowest SBP and better aortic compliance than cluster 1, suggesting a relatively favourable vascular profile (26). However, they carried an intermediate risk of MACE, particularly for AF and all-cause mortality, as shown in sensitivity analyses. This risk profile aligns with evidence that MetS promotes AF through atrial remodelling (both structural and electrical), cardiac autonomic changes, and systemic inflammation (27,28). Women are more vulnerable to MetS-related CV risk due to genetic predisposition, postmenopausal metabolic shifts, and sociocultural factors, as seen in this study (28,29). While obesity, diabetes, and dyslipidaemia can each contribute to cardiac remodelling, their coexistence can amplify these effects, markedly increasing AF susceptibility (27,28). This may explain the elevated AF risk in this female-dominant group, where multiple MetS components were present.

Interestingly, despite significant LA dysfunction in both cluster 2 and 3, neither showed increased LAVi. This highlights the heterogeneous nature of LA response to HTN and suggests that functional deterioration may precede overt LA dilatation or even LV hypertrophy, as observed in cluster 3. This emphasises the value of assessing LA function as an earlier predictor of AF risk in hypertensive patients, particularly those who do not yet exhibit LV hypertrophy, rather than relying solely on LA size (30–34).

The sensitivity analysis showed that including subjects with subclinical cardiomyopathies did not significantly change the clustering results but slightly overestimated HF risk and the mediating role of LV hypertrophy in death risk for cluster 2. In cluster 3, their inclusion attenuated the death risk and the roles of atrial function and LV GLS in risk mediation. Overall, these findings confirm that clustering-derived risk profiles are robust and accurately reflect underlying phenotypes.

### Imaging insights into pathophysiological mechanisms of risks

Analysing the imaging findings in relation to outcomes provided key insights into the pathophysiological mechanisms underlying each hypertensive phenotype. Clusters in the order 1, 3, and 2 showed a stepwise increase in concentric LV hypertrophy, worsening LV mechanics, and declining diastolic function (lower LA expansion index), mirroring their escalating risk of adverse events. While these changes align with hypertensive heart disease progression, the interplay of comorbidities uniquely shaped these patterns (35–37).

The significant LV remodelling and global impairment in LV mechanics (including GLS, GCS, and GRS) observed in cluster 2 resemble the HF with preserved ejection fraction substrate (9). The LA dysfunction, particularly in reservoir and conduit function, is primarily driven by the high comorbidity burden, especially prevalent MI and systemic organ dysfunction, which further increases CV risk (25,33,34,38,39).

The cardiac changes observed in cluster 3, featuring less severe LV remodelling yet LA dysfunction across all phases (reservoir, conduit, and contractile), likely reflect the metabolic and inflammatory burden of MetS, which promotes atrial remodelling prior to the emergence of significant LV changes, thus creating an arrhythmic substrate for AF risk (30–34,37).

Similar patterns of LA dysfunction have been reported in specific HTN populations and in cohorts susceptible to AF with elevated metabolic risk factors (14,32,34). This underscores how MetS-induced systemic pathways contribute to atrial remodelling, thereby increasing AF susceptibility.

Both clusters 2 and 3 showed an increased AF risk, confirming the well-established bidirectional HTN-AF connection, though driven by two distinct mechanisms (40). In cluster 2, HTN-induced remodelling and coexisting atherosclerosis created a pro-HF environment that heightened AF susceptibility, reinforcing evidence that HF with preserved ejection fraction and AF share a common myocardial disease substrate (41). Conversely, in cluster 3, MetS-related metabolic and inflammatory factors primarily induced atrial changes that made individuals more prone to AF, emphasising the necessity for targeted risk-factor management in this group (14,34).

The mediation analyses further clarified how these phenotypic differences influenced adverse events. In cluster 2, where LV remodelling was most pronounced, LV hypertrophy and LV mechanical dysfunction emerged as key mediators of clinical risk, especially for HF. LA dysfunction, instead, played a greater role in all-cause mortality. These findings confirm the prognostic impact of LV hypertrophy in hypertensive heart disease (40,42). In cluster 3, outcomes were generally less influenced by LV structures and more driven by LA dysfunction, reinforcing a metabolic-inflammatory route to atrial remodelling, which conventional CMR metrics may not fully capture.

Viewed across the spectrum of hypertensive heart disease, these three phenotypes represent distinct patterns of cardiac and vascular involvement, shaped by the interplay of elevated afterload, metabolic factors, and comorbidities. Cluster 1 represents a milder form with minimal CV remodelling and a more favourable prognosis. Cluster 3 represents an intermediate-risk phenotype dominated by MetS-driven diastolic dysfunction and moderate CV risk, especially for AF. Cluster 2 reflects a more advanced stage, where severe LV remodelling and vascular dysfunction converge, approaching the HF phenotype and carrying the highest risk of adverse outcomes. These phenotypes represent distinct, rather than strictly progressive, profiles that may inform tailored therapeutic strategies.

### Comparison with previous studies using clustering in hypertension

Several studies have previously applied clustering to hypertensive populations, identifying clinically relevant phenotypes. Yang et al., using hierarchical clustering in the SPRINT trial cohort (n=9,361), identified four groups, including an “extra-risky” subset resembling cluster 2, and an “obese” subset similar to cluster 3 (5). However, the exclusion of diabetic patients in their study, limiting direct comparability. Guo et al. applied K-means clustering to essential HTN patients (n=513), finding groups approximating cluster 3 (female, diabetic) and cluster 2 (high prevalence of coronary artery disease), though they did not link the phenotypes to incident events (6).

Vaura et al. (n=3,726; FINRISK cohorts) and Bala et al. (n=698; Romanian population-based study) described high-risk metabolic profiles in both sexes (e.g., high body mass index and glucose level), confirming the synergistic effects of metabolic dysfunction and HTN on CV risk (7,8). Unlike these studies, which identified a single high-risk metabolic profile, this work revealed two distinct high-risk pathways: atherosclerosis-driven (cluster 2) and metabolic-driven (cluster 3). The ability of this study to discern better risk profiles likely stems from the larger sample size and the greater number of clinical variables used for clustering.

Katz et al. (2017) (n=1,273, HyperGEN) integrated echocardiography with clinical metrics to identify two distinct CV phenotypes of HTN. They found a high-risk group resembling cluster 2 and sharing characteristics with HFpEF (9). However, they did not relate phenotypes to incident events or identify an intermediate CV phenotype like cluster 3.

Although these previous studies varied in terms of populations, clustering variables, and algorithms used, they generally align with the current findings, emphasising that factors such as atherosclerotic burden and metabolic disorders shape cardiovascular risk, beyond blood pressure alone.

### Clinical implications

Identifying distinct HTN phenotypes can enable tailoring prevention and management strategies. For individuals resembling cluster 1, interventions should promote a healthy lifestyle and control blood pressure to prevent disease progression. In contrast, cluster 3 individuals, despite lower SBP, present a high metabolic and inflammatory burden, necessitating interventions beyond blood pressure targets. This involves tackling obesity, insulin resistance, and dyslipidaemia, as well as using medications aimed at improving diastolic dysfunction to reduce the risk of AF (29). Cluster 2, instead, characterised by advanced atherosclerosis and target organ disease, requires a more intensive approach, including strict blood pressure (including DBP) and lipid control, medications to counteract LV remodelling and improve microvasculature, and targeted lifestyle modifications to lower CV risk.

AF prevention should align with each phenotype’s underlying mechanisms. In cluster 2, limiting HF progression and LA remodelling is key to reducing the arrhythmogenic substrate. Conversely, in cluster 3, intensive metabolic and inflammatory control is paramount, as evidence suggests optimal metabolic management significantly lowers AF incidence in HTN (29).

### Strengths and limitations

A key strength of this study is its large sample size, and the extensive clinical data used for clustering. Unlike most prior clustering research, which has explored cluster relevance through either clinical outcomes or imaging findings alone, this study integrates both, offering deeper insights into hypertensive heart disease risk pathways.

This study does not aim to redefine hypertension classification, as the identified clusters may vary depending on population characteristics and data availability. Rather, it highlights the potential of a multidimensional, data-driven approach to uncover clinically relevant phenogroups that could inform tailored interventions. Nevertheless, these findings align with other clustering studies in hypertensive cohorts, supporting their broader clinical relevance. To move beyond proof-of-concept, however, external validation in independent datasets and prospective comparisons with traditional risk tools are necessary to confirm their added clinical utility.

Several limitations related to the dataset used should be noted. The cohort is predominantly middle-aged and Caucasian, limiting generalisability to other populations. Self-reported data (e.g. lifestyle, conditions) may be biased, leading to some inaccuracy. Detailed information on medication classes, dosages, and durations (including antihypertensive, lipid-lowering, and other relevant therapies) was not available for this work and may have influenced the observed phenotypic patterns. Genetic data from the UK Biobank were not integrated, which could have enhanced risk stratification and will be explored in future work. Future work will aim to integrate both genetic and medication data to improve clustering precision and assess whether phenotype-guided strategies can support more tailored interventions and improve long-term cardiovascular outcomes.

Despite greater remodelling and presumed fibrosis, clusters 2 and 3 exhibited lower native T1 values than cluster 1, aligning with prior UK Biobank findings yet conflicting with other studies (22,43). Variations in acquisition methods or complex fibrotic patterns within HTN, which perhaps native T1 alone cannot fully capture, may account for these discrepancies (44,45). Incorporating contrast-enhanced imaging and extracellular volume quantification, which are unavailable in this dataset, could enhance the assessment of fibrotic patterns in HTN and clarify such discrepancies (45).

Finally, although *post hoc* analyses and bootstrapping supported the clinical significance and confirmed stability of the clustering method, replicating these findings in independent cohorts is essential to confirm their generalizability.

## Conclusions

Unsupervised clustering of clinically available data in a large UK Biobank cohort of HTN subjects identified three distinct phenotypes with differing risk profiles, CV imaging characteristics, and outcomes. These phenotypes likely reflect different underlying mechanisms and support a more individualised approach to risk assessment. These findings underscore the heterogeneity of hypertensive heart disease and demonstrate the potential of data-driven strategies to complement traditional risk stratification and guide personalised care.

## Data availability statement

This research was conducted using the UK Biobank resource under access application 2964. UK Biobank will make the data available to all bona fide researchers for all types of health-related research that is in the public interest, without preferential or exclusive access for any persons. All researchers will be subject to the same application process and approval criteria as specified by UK Biobank. For more details on the access procedure, see the UK Biobank website: http://www.ukbiobank.ac.uk/register-apply/.

## Author contributions

ER conceived the idea, conducted the analyses, and wrote the manuscript. GGS, NA, and SEP supervised and guided the experiments. SC, HN, PBM, and AM provided feedback on the clinical interpretation and revised the manuscript. AMS, MA, and CRSB advised on the machine learning analysis, and JC advised on statistics. All the authors contributed to the work and read and approved the final manuscript.

## Abbreviations

AF: Atrial fibrillation
ARI: Adjusted rand index
CV: Cardiovascular
DBP: Diastolic blood pressure
FAMD: Factorial analysis of mixed data
GCS: Global circumferential strain
GLS: Global longitudinal strain
GRS: Global radial strain
HDL: High-density lipoprotein
HF: Heart failure
HR: Hazard ratio
HTN: Hypertension
LA: Left atrial
LAEF: Left atrial emptying fraction
LAV: Left atrial volume
LDL: Low-density lipoprotein
LV: Left ventricle
MACE: Major adverse cardiovascular events
MetS: Metabolic syndrome
MI: Myocardial infarction
M/V: Mass-to-volume
RV: Right ventricle
SBP: Systolic blood pressure
SHAP: Shapley additive explanations

